# Celiac Disease Risk Allele Frequencies in San Luis (Argentina) and Evaluation of a Saliva Direct PCR Genotyping Approach

**DOI:** 10.64898/2026.05.19.26353109

**Authors:** Celia Pérez, Constanza Pistone, Camila Romero, Angeles Carrillo, Jimena Manzur, Constanza Chialva, Hector Quiroz, Maximiliano Juri Ayub

## Abstract

Celiac disease (CD) is strongly associated with specific HLA-DQ heterodimers, formed by HLA-DQA1 and HLA-DQB1 proteins. In particular DQ2.5 (DQB1*02 associated to DQA1*05) and DQ8 (DQB1*03:02 with DQA1*03) are present in virtually all celiac patients. HLA-DQB1*02 is considered the main single genetic susceptibility marker and has been reported in 90–95% of CD patients. However, the distribution of these alleles may vary across populations, potentially impacting the performance of genetic screening strategies. In this study, we evaluated the prevalence of HLA-DQ2.5 and DQ8 genotypes in celiac patients (n = 41) and an unbiased general population cohort (n = 60) from San Luis, Argentina, using a PCR-based genotyping approach. In addition, we assessed the feasibility of a simplified saliva direct PCR protocol for large-scale testing. Overall, 95.1% of CD patients carried DQ2.5 and/or DQ8. Notably, 41.5% of patients were DQ8(+)/DQ2.5(−), and 36.6% lacked the DQB1*02 allele, indicating that DQB1*02-based screening alone would have reduced sensitivity in this population. In the general population, 53.3% of individuals carried CD-associated genotypes, with a markedly higher prevalence of DQ8 compared to European cohorts. Genotype distributions deviated from Hardy–Weinberg equilibrium in CD patients but not in the general population. We show that DQB1*03:02 is a reliable proxy for DQ8, allowing simplification of genotyping strategies, whereas DQA1*05 typing remains essential to discriminate DQ2.5 from other lower-risk DQB1*02 carrying heterodimers. We also describe a saliva direct PCR approach showing a performance comparable to purified DNA-based assays. These findings highlight the importance of population-specific genetic data for optimizing CD screening strategies and foster the development of simplified, cost-effective genotyping approaches for large-scale applications.

## INTRODUCTION

Celiac disease (CD) is an immune disorder triggered by gluten ingestion in genetically predisposed individuals [1]. Four alleles at two human leukocyte antigen (HLA) loci (DQA1*03 and *05, and DQB1*02 and *03:02) are known to be associated with CD. Moreover, the absence of these alleles has a high negative predictive value. The heterodimers DQ2.5 (DQB1*02 associated to DQA1*05) and DQ8 (DQB1*03:02 associated to DQA1*03) significantly increase the risk of developing CD.

HLA-DQB1*02 is currently considered the strongest predictor of genetic susceptibility and has been reported to be present in 90–95% of patients with CD (Poddighe et al., 2020), although this frequency may vary across populations. Therefore, it has been suggested that large-scale genetic testing for this allele would be desirable if cost-effective strategies were available [2–6]. Moreover, a two-step screening strategy has been proposed for children: first-line screening for HLA-DQB1*02, followed by prospective serological studies focused on DQB1*02-positive individuals [7]. Recently, we reported a direct saliva-based real-time PCR protocol for detecting the presence of HLA-DQB1*02 [8].

Most PCR-based assays for HLA typing (particularly HLA-DQ2.5 and HLA-DQ8) have been traditionally validated using genomic DNA obtained from peripheral blood, which requires invasive sampling, trained personnel, and appropriate biosafety conditions. However, saliva is a reliable alternative source of genomic DNA, enabling simple self-collection by patients without the need for clinical intervention, while maintaining sufficient DNA yield and quality for molecular analyses [9, 10]. In addition, saliva offers important logistical advantages, as DNA in these samples is relatively stable and can be transported under simple refrigeration conditions without additives. Finally, saliva self-collection is a non-invasive approach that improves patient compliance and is particularly advantageous in pediatric populations or in individuals with needle aversion. Collectively, these features position saliva as an attractive alternative for the development of more accessible and scalable genotyping strategies in the context of celiac disease.

In the present work, we initially used DNA purified from saliva along with PCR-based genotyping to evaluate the prevalence of DQ2.5 and DQ8 in both celiac patients and the general population from the city of San Luis and surrounding areas. We tested for the presence of the alleles HLA-DQA1*03*/*05 and HLA-DQB1*02*/*03:02. Based on these results, genotypes were classified according to the aility to yield risk heterodimers, as indicated in **Table 1**. Relative frequencies of alleles and DQ heterodimers were analyzed and compared with previously reported data, and the relevance of local population characteristics is discussed. In addition, as a proof of concept, we explored the efficiency of a simplified, even lower cost genotyping strategy using direct saliva PCR, omitting the DNA purification step, with promising results.

**Table 1.** DQA1*X and DQB1*X indicate alleles different from DQA1*03/05 and DQB1*02/0202, respectively.

|  |  | DQB1 | DQA1 |
| --- | --- | --- | --- |
| Risk DQ | DQ2.5 | *02 | *05 |
|  | DQ2.X |  | No *05 |
|  | DQ8 | *0302 | *03 |
| Low Risk DQ | Negative | *X | *03/05/X |

## METHODS

### Samples and DNA purification

Saliva samples were obtained from 41 volunteers with confirmed CD. All participants provided written informed consent, and the study protocol was approved by the Ethics Committee of the Health Ministry of San Luis Province. To obtain an unbiased set of nucleic acid samples representative of the general population from the same geographical region, remnants of 60 purified samples from oro/nasopharyngeal swabs were obtained from the Public Health Laboratory of San Luis Province. These samples were received in an irreversibly anonymized form, in accordance with approval from the corresponding Ethics Committee. Saliva samples were used both for DNA purification, as previously described [8], and/or directly as PCR templates.

### Genotyping PCR strategy

Genotyping was performed using two duplex real-time PCR assays targeting HLA-DQA1 and HLA-DQB1, respectively. Each assay amplified a region containing a polymorphic site interrogated by two alternative allele-specific hydrolysis (*TaqMan*) probes labeled with distinct fluorophores. The combination of fluorescence signals generated in each reaction enabled discrimination among allelic variants, as summarized in **Table 2**. Primer and probe sequences are not disclosed due to proprietary restrictions.

**Table 2.**
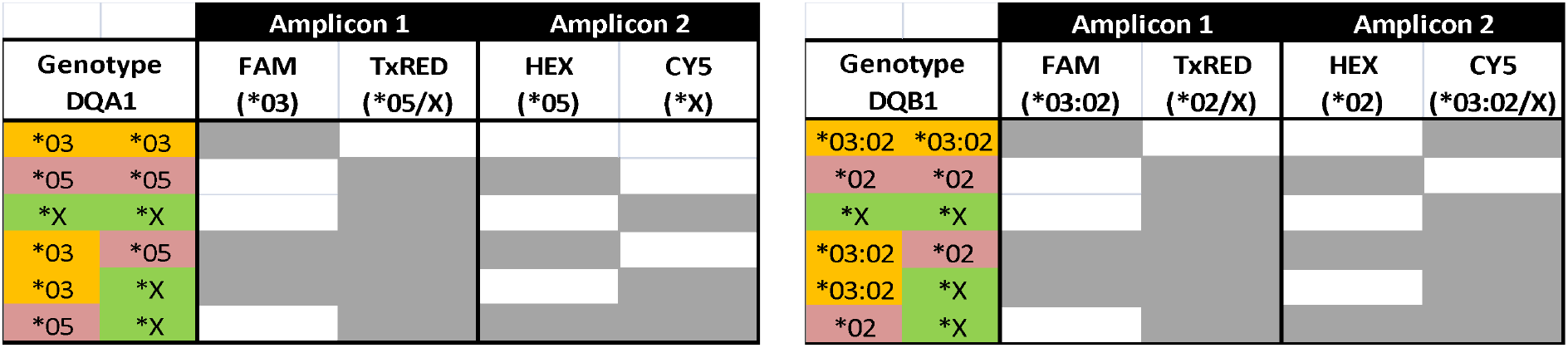
Genotypes for HLA-DQA1 (left) and HLA-DQB1 (right), with their corresponding probe signal patterns. Positive amplification signals are shown in grey and negative signals in white for each probe.

When purified DNA was used as template, *Taq* DNA polymerase (Inbio Highway, Kit T-Plus Free ADN Polimerasa), in the presence of MgCl_2_ 5 mM was employed. For the saliva direct protocol, 5 µL of saliva diluted 1:10 in nuclease-free water was used as template, in combination with LightCycler® Multiplex RNA Virus Master (Roche), following the manufacturer’s instructions, except that addition of reverse transcriptase was omitted.

Real-time PCR cycling conditions were as follows: initial denaturation at 95°C for 3 min; 10 initial cycles of 60°C for 30 s and 95°C for 30 s without fluorescence acquisition; followed by 35 amplification cycles of 60°C for 30 s and 95°C for 30 s, with fluorescence acquisition at both temperatures. When indicated, a final melting curve analysis was performed from 60°C to 98°C with continuous fluorescence acquisition.

### Sanger sequencing genotyping

To validate the PCR-based genotyping, 30 samples from the general population were analyzed by Sanger sequencing. Primers used for the amplification of HLA-DQA1 and HLA-DQB1 are listed in **Table 3**. Two different amplicons were used for HLA-DQA1 to mitigate allele dropout and to account for the presence of indels in some alleles that may impair sequence interpretation. Genotypes assigned by PCR were 100% coincident with Sanger sequencing results.

**Table 3.**
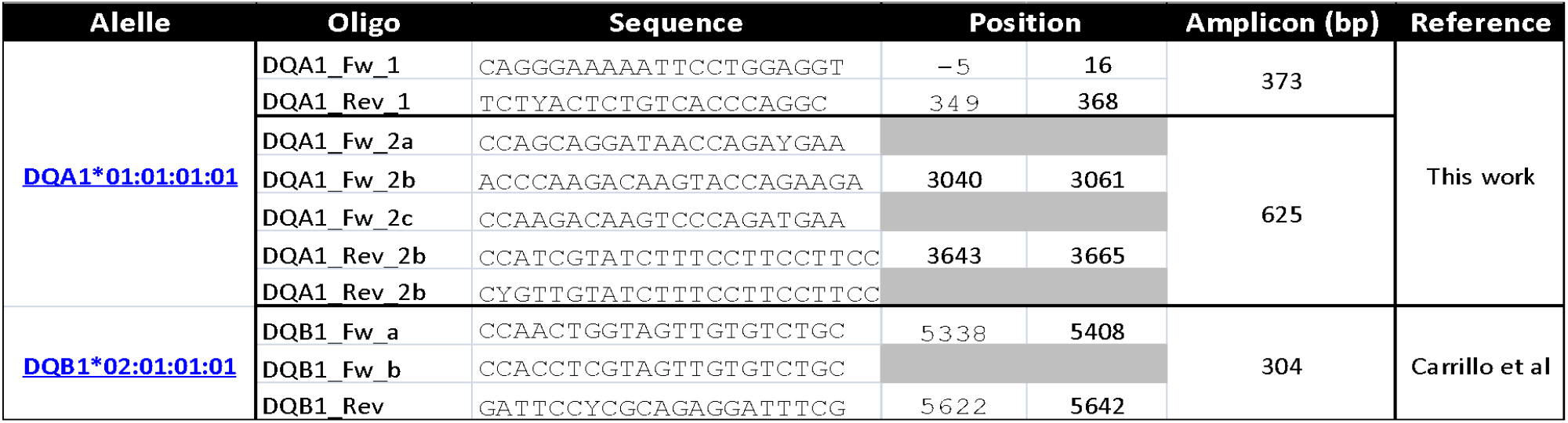
Primers used for PCR amplification and Sanger sequencing of HLA-DQA1 and HLA-DQB1 *loci* for genotype assignment. All oligonucleotides are shown relative to the reference sequences indicated in the left column, with amplicon sizes provided in base pairs. PCR products were generated and subsequently sequenced by Sanger to determine allelic variants. For some targets, multiple forward and/or reverse primers were included due to sequence polymorphisms among alleles, ensuring robust amplification across different genotypes. Grey-shaded cells indicate primers that do not anneal to the reference sequence shown but instead bind to the corresponding regions in alternative alleles; therefore, position numbers are not provided. Reference sequences are available at https://www.ebi.ac.uk/ipd/imgt/hla/alleles.

## RESULTS

### 1. Prevalence of HLA-DQ2.5 and DQ8 heterodimers in celiac patients

Saliva samples from patients with confirmed celiac disease (CD; n = 41) were used for DNA purification and genotyped by a real-time PCR protocol based on allele-specific hydrolysis probes. Detailed genotyping results are provided in Supplementary Table 1. Overall, 95.1% (**IC**_**95**_ 83.5-99.4%) of samples (39/41) were positive for DQ2.5 (DQB1*02/DQA1*05) and/or DQ8 (DQB1*03:02/DQA1*03), in agreement with previous reports. One sample (2.4%; **IC**_**95**_ 0-12.8%) was classified as DQ2.X (DQB1*02/DQA1*X), and one sample (2.4%; **IC**_**95**_ 0-12.8%) lacked all risk alleles (DQB1*X/DQA1*X). These values are consistent with those reported in other populations ([11] and references 26-30 herein).

Notably, 41.5% (**IC**_**95**_ 26.3-57.9%) of CD patients (17/41) were DQ8(+)/DQ2.5(−), a proportion markedly higher than the <10% typically reported in European cohorts [11]. In addition, 36.6% (**IC**_**95**_ 22.1-53.1%) of patients (15/41) lacked the DQB1*02 allele. These findings indicate that screening strategies based solely on DQB1*02 detection [12], although cost-effective in other regions, would have limited sensitivity in our population.

### 2. High prevalence of DQ8 in local population

We next analyzed an unbiased set of DNA samples from the general population (GP; n = 60). Overall, 53.3% (**IC**_**95**_ 40.0-66.3%) of individuals (32/60) carried alleles encoding DQ2.5 and/or DQ8 heterodimers (Supplementary Table 2), while an additional 10% harbored HLA-DQB1*02 without DQA1*05 (yielding DQ2.X heterodimers). Consequently, fewer than 40% of individuals lacked CD-associated risk genotypes. In comparison, approximately 70% of individuals in European populations are reported to lack these genotypes [3]. While the prevalence of DQ2.5(+) genotypes was similar to previous reports, the frequency of DQ8(+) genotypes in our population (30%) was substantially higher than that reported in European cohorts (~7.4%). These data suggest that the increased prevalence of DQ8 in local CD cohort is primarily explained by its higher baseline prevalence in the general population.

Figure 1 summarizes the distribution of HLA-DQ heterodimers in both CD patients and the general population.

**Figure 1.**
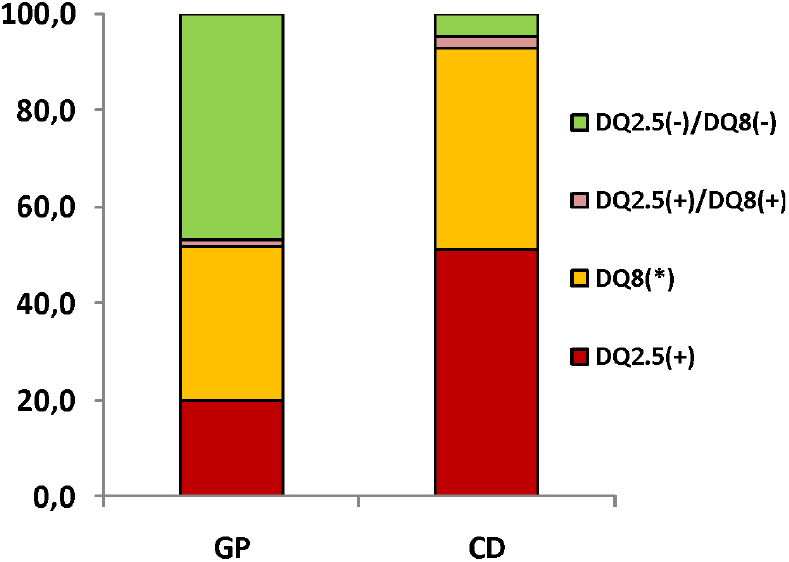
Percent of DQ heterodimers in general population (GP) and celiac population (CD).

### 3. DQB1 alleles as proxies of DQ heterodimers

In the general population cohort, 35% (**IC**_**95**_ 23.1-48.4) of individuals (21/60) carried the DQB1*02 allele. Among them, 61.9% (13/21) also carried DQA1*05 and 38.1% (8/21) were DQA1*05(-). This indicates that DQB1*02 is not a reliable proxy for the DQ2.5 heterodimer. In contrast, 33.3% (**IC**_**95**_ 21.7-46-7%) of individuals (20/60) carried DQB1*03:02, and all of them were also positive for DQA1*03. Thus, DQB1*03:02 showed complete concordance with DQ8, making it an excellent proxy for this heterodimer. Similar results have been observed in other cohorts by Megiorni *et al* [3]. These results suggest that DQA1*03 genotyping could be omitted without loss of information, whereas DQA1*05 genotyping provides important discrimination between DQ2.5 and DQ2.X genotypes.

### 4. Relative risk calculation based on genotypes frequencies

Genotype frequencies in CD patients and the general population were used to estimate relative risk (RR), using the absence of risk alleles (NEG) as the reference category (RR=1). As expected, the highest risk was associated with the DQ2.5 genotype (RR = 38.5). The presence of DQB1*02 alone (without DQA1 information) was also associated with a high relative risk (RR = 26.9). Importantly, inclusion of DQA1*05 genotyping allowed discrimination between DQ2.5 (RR = 38.5) and DQ2.X (RR = 3.7), highlighting its added value. Consistent with the complete linkage observed, DQB1*03:02 can be considered equivalent to DQ8 for risk estimation purposes (RR = 18.1). **Figure 2** shows the relative risk associated with each genotype category.

**Figure 2.**
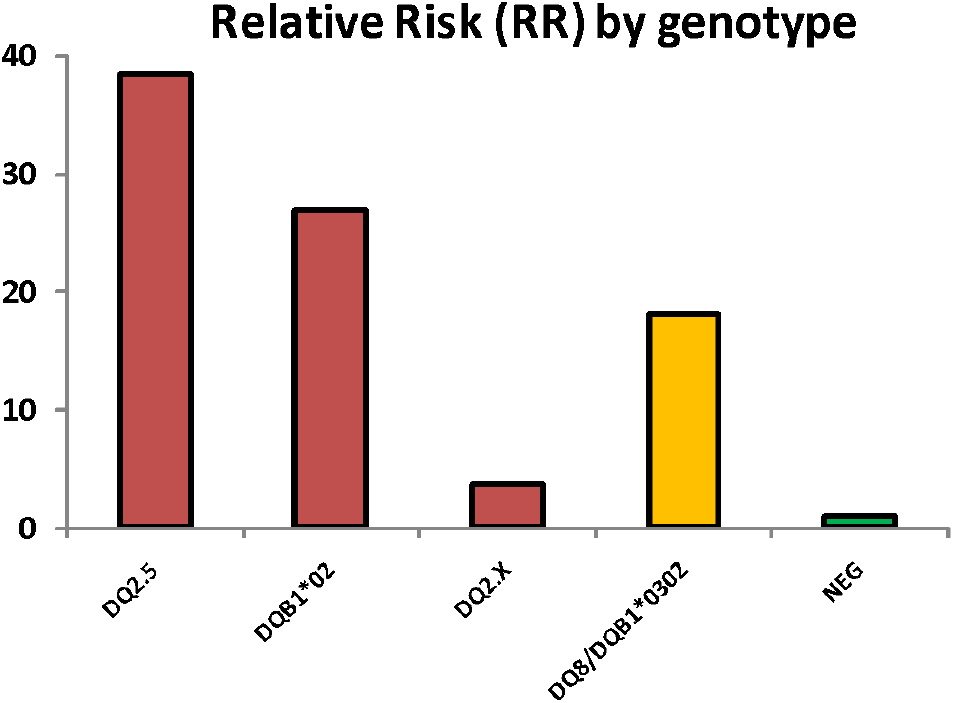
Relative risk of Celiac Disease associated to different genotypes. The ratios of genotypes frequencies in CD and general population were calculated and normalized to negative genotype (lacking of risk alleles) as unit reference (green). Genotypes carrying DQB1*02 (red) and DQB1*03:02 (yellow) are indicated.

### 5. Hardy-Weinberg analyses

Using genotype data from **Supplementary Tables 1** and **2**, we evaluated whether the observed genotype frequencies were consistent with Hardy–Weinberg equilibrium expectations. As shown in **Tables 4** and **5**, genotype distributions for both HLA-DQA1 and HLA-DQB1 in the CD cohort showed significant departures from equilibrium (χ^2^ test, p < 0.05), whereas no significant deviations were detected in the general population.

**Table 4.** Observed and expected genotype count for DQA1 and DQB1 alleles in celiac and general cohorts. Chi^2^ values are calculated for each genotype.

| General population |  |  |  |  |
| --- | --- | --- | --- | --- |
| DQA1 |  | Observed | Expected | Chi<sup>2</sup> |
| *05 | *05 | 5 | 3,27 | 0,92 |
| *03 | *03 | 2 | 4,27 | 1,20 |
| *X | *X | 14 | 15,00 | 0,07 |
| *05 | *03 | 7 | 7,47 | 0,03 |
| *05 | *X | 15 | 14,00 | 0,07 |
| *03 | *X | 17 | 16,00 | 0,06 |
|  |  | 60 | 60,00 | 2,35 |
| DQB1 |  | Observed | Expected | Chi<sup>2</sup> |
| *02 | *02 | 3 | 2,40 | 0,15 |
| *03:II | *03:II | 2 | 2,02 | 0,00 |
| *X | *X | 22 | 22,82 | 0,03 |
| *02 | *03:II | 3 | 4,40 | 0,45 |
| *02 | *X | 15 | 14,80 | 0,00 |
| *03:II | *X | 15 | 13,57 | 0,15 |
|  |  | 60 | 60,00 | 0,78 |

| Celiac population |  |  |  |  |
| --- | --- | --- | --- | --- |
| DQA1 |  | Observed | Expected | Chi<sup>2</sup> |
| *05 | *05 | 5 | 3,81 | 0,37 |
| *03 | *03 | 6 | 5,49 | 0,05 |
| *X | *X | 2 | 4,45 | 1,34 |
| *05 | *03 | 5 | 9,15 | 1,88 |
| *05 | *X | 15 | 8,23 | 5,57 |
| *03 | *X | 8 | 9,88 | 0,36 |
|  |  | 41 | 41,00 | 9,57 |
| DQB1 |  | Observed | Expected | Chi<sup>2</sup> |
| *02 | *02 | 5 | 5,86 | 0,13 |
| *03:II | *03:II | 5 | 3,23 | 0,98 |
| *X | *X | 1 | 4,78 | 2,99 |
| *02 | *03:II | 4 | 8,70 | 2,54 |
| *02 | *X | 17 | 10,59 | 3,89 |
| *03:II | *X | 9 | 7,85 | 0,17 |
|  |  | 41 | 41,00 | 10,68 |

**Table 5.** Chi^2^ total values for each gene (DQA1 and DQB1) in celiac and general cohorts are shown along with their respective significance p values.

|  | General population |  | Celiac population |  |
| --- | --- | --- | --- | --- |
|  | DQA1 | DQB1 | DQA1 | DQB1 |
| **Chi<sup>2</sup>** | 2,35 | 0,78 | 9,57 | 10,68 |
| **p** | 0,503 | 0,85 | 0,022 | 0,013 |

### 6. Saliva direct PCR

We previously reported a saliva direct real-time PCR protocol for DQB1*02 genotyping based on melting temperature differences associated with a allele-specifc 52 bp insertion [8]. As can be deduced from our genotyping local data, DQB1*02 information is relevant but not enough for a accurate relative risk assignation, since DQB1*03:02 and DQA1*05 add significant information to this goal.

Then, we explored the feasibility of adapting our probe-based assays to a saliva direct format. Given that DQB1*03:02 showed complete concordance with DQ8, DQA1*03 detection was omitted in the saliva direct format. Instead, DQA1*05 detection was retained, as it is required to discriminate between DQ2.5 (high risk) and DQ2.X (low risk). Under this simplified design, DQA1*03/03 samples are expected to produce no probe signal, making them indistinguishable from amplification failure (see **Table 2**). To address this limitation, amplification of the second DQA1 product was performed in the presence of the intercalating dye EvaGreen, allowing confirmation of amplification by melting curve analysis. To prevent interference of EvaGreen with HEX-probe fluorescence, amplification signal was acquired at 95°C, where DNA is fully denatured. In this way, DQA1*05 positive samples are detected by HEX signal, whereas DQA1*05 negative samples are expected to yield negative HEX signal and a PCR product with a Tm of around 78ºC. **Supplementary Figure 1** shows the main similarities and differences between the DNA and saliva direct based PCR.

Different enzymes and commercial qPCR mixes were evaluated for the DQA1*05 singleplex and DQB1*02/03:02 duplex reactions using crude saliva as template. The best performance was obtained with LightCycler® Multiplex RNA Virus Master (Roche). A set of 20 samples with representative diversity of genotypes was analyzed as proof of concept. **Figure 3** shows representative results from saliva direct protocol for DQ2.5(+) and DQ8(+). A complete set of genotypes can be seen in **Supplementary Figure 2**.

**Figure 3.**
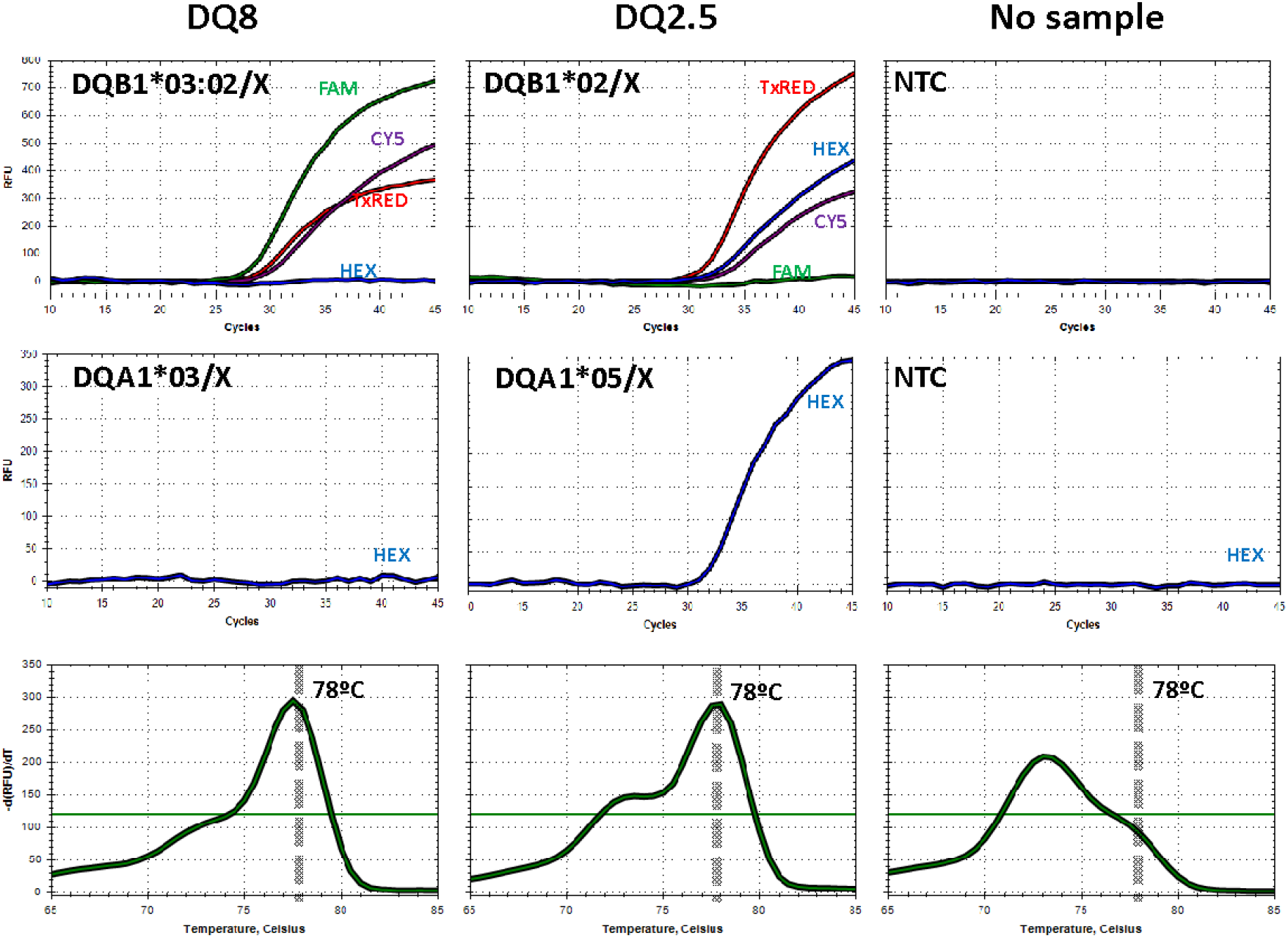
Representative results of genotyping by real-time PCR using the saliva direct format. **Upper panel**. Duplex HLA-DQB1 assay with allele-specific hydrolysis probes for the detection of DQB1*02, DQB1*03:02 and DQB1*X. **Middle panel**. Singleplex PCR for detection of the HLA-DQA1*05 allele based on a specific HEX-labelled probe. **Lower panel**. Melting curve of the HLA-DQA1 amplicon to confirm successful amplification. Samples lacking DQA1*05 are identified by a negative HEX signal combined with a specific melting peak (Tm=78ºC), thereby ruling out false negative results.

## DISCUSSION

Our results confirm the strong association between HLA-DQ heterodimers (DQ2.5 and DQ8) and celiac disease, but also reveal substantial population-specific differences that have important implications for genetic screening strategies. While the overall proportion of CD patients carrying DQ2.5 and/or DQ8 was consistent with previous reports, the markedly higher prevalence of DQ8 challenges the proposal of DQB1*02 alone as an adequate first-line screening marker [4–6].

In this context, strategies based exclusively on DQB1*02 detection, although effective in European populations, would show limited sensitivity in our setting. Instead, our data support the inclusion of DQB1*03:02 and DQA1*05 genotyping to improve risk stratification. The complete concordance observed between DQB1*03:02 and DQ8 suggests that simplified assays can omit DQA1*03 without loss of information, whereas DQA1*05 remains critical to distinguish high-risk (DQ2.5) from lower-risk (DQ2.X) genotypes.

From a methodological perspective, we demonstrate the feasibility of adapting probe-based genotyping assays to a saliva direct format, which eliminates the need for DNA purification and reduces cost and time for testing. However, this simplification introduces a key limitation: the absence of probe signal in certain genotypes cannot be distinguished from amplification failure. We address this by incorporating melting curve analysis using an intercalating dye, providing an internal control of amplification.

The use of direct saliva as input material emerges as a promising alternative for molecular diagnostics, particularly due to its non-invasive nature, ease of collection, and reduced need for sample processing. While further optimization and validation may be required before routine implementation in clinical diagnostic settings, the current protocol already demonstrates robustness and practicality for large-scale applications. In this context, its simplicity, low cost, and compatibility with high-throughput workflows make it especially suitable for epidemiological and population-based studies, where rapid and accessible genotyping strategies are essential.

An important feature of our genotyping strategy, particularly relevant in the saliva-direct format, is its robustness against false-negative results. The same PCR product serves both as the diagnostic target (via hydrolysis of allele-specific probes) and as an internal control (either through hydrolysis of at least one of two alternative probes or by melting curve analysis). In this way, true negative results can be distinguished from false negatives based on the presence or absence of PCR amplification, respectively.

Finally, the successful implementation of direct saliva-based genotyping for celiac disease constitutes a proof of concept that can be readily extended to other diagnostic settings requiring the detection of specific genetic polymorphisms.

## Data Availability

All data produced in the present study are available upon reasonable request to the authors

## ACKNOWLEDGEMENTS

We are deeply indebted to the volunteers who participated in this study and to Melisa Leyes for collecting part of the saliva samples. We also thank Eliana Rosales (Provincial Public Health Laboratory “Dalmiro Pérez Laborda”) for providing anonymized nucleic acid samples. CP, JM, and MJA are a postdoctoral fellow, a support professional, and an investigator at CONICET, respectively, and also serve as professors at the Universidad Nacional de San Luis (UNSL). CR and AC are research interns at the LDDM, UNSL. Specific primers and probes were provided by Biocientífica S.A. This work was supported by grants from UNSL (PROICO 02-1720) and CONICET (PIP 2021–2023 No. 2732).

---

**Supplementary Figure 1.**
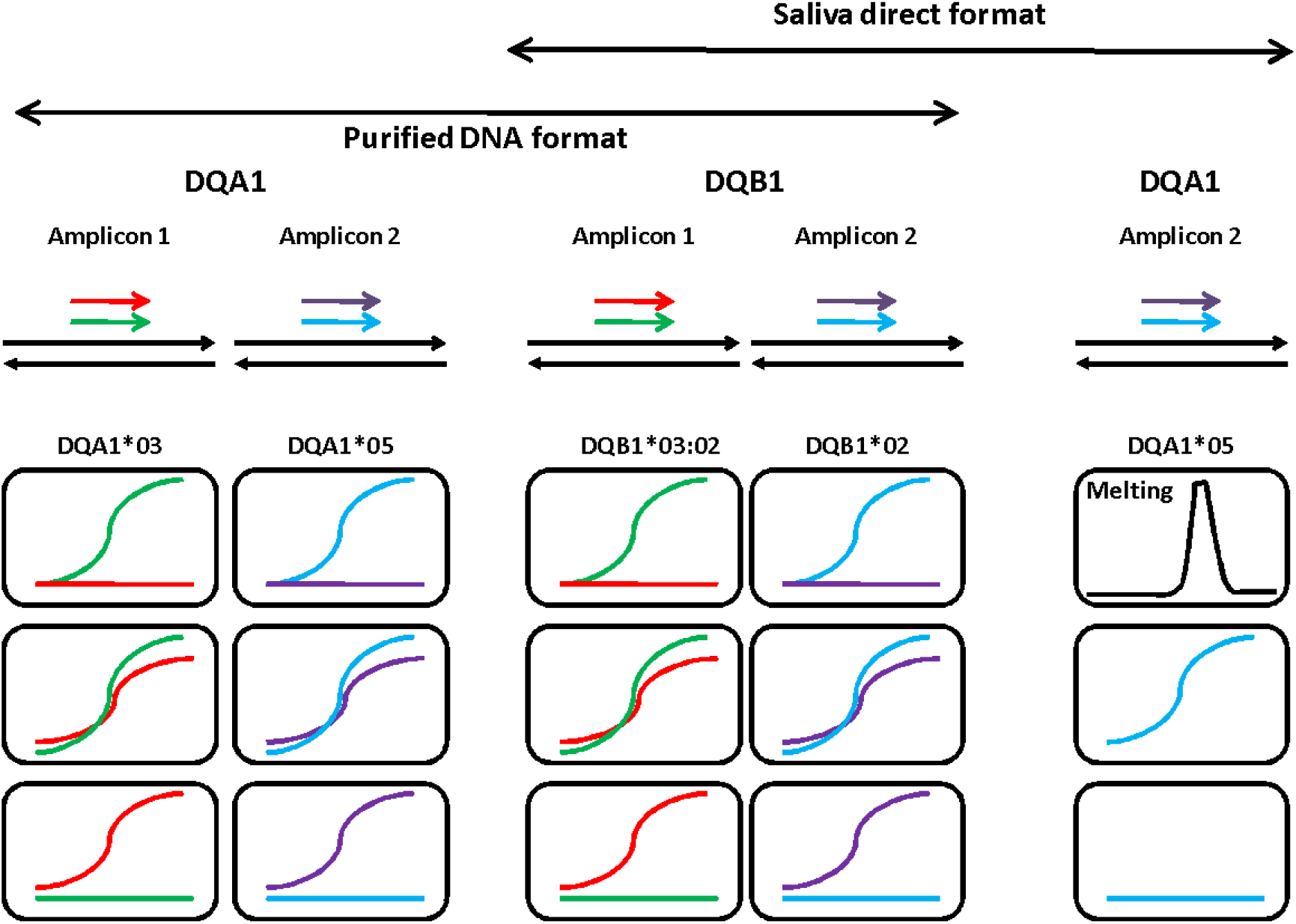
Schematic comparison between purified DNA-based and saliva direct real-time PCR formats for HLA-DQA1 and HLA-DQB1 genotyping. In both settings, genotyping relies on allele-specific hydrolysis probes targeting HLA-DQB1*02/03:02 and HLA-DQA1*05, enabling identification of DQ2.5, DQ2.X and DQ8-associated genotypes. In the purified DNA format, amplification specificity and signal interpretation are based solely on probe fluorescence patterns. In the saliva direct format, the assay is simplified by omitting DQA1*03 detection, as DQB1*03:02 fully predicts DQ8. Under this design, samples lacking DQA1*05 do not generate probe signal, making them indistinguishable from amplification failure based on fluorescence alone. To overcome this limitation, successful amplification is monitored using an intercalating dye, allowing melting curve analysis. Thus, while both formats share the same probe-based genotyping basis, the saliva direct approach requires an additional melting step to validate amplification in probe-negative samples.

**Supplementary Figure 2.**
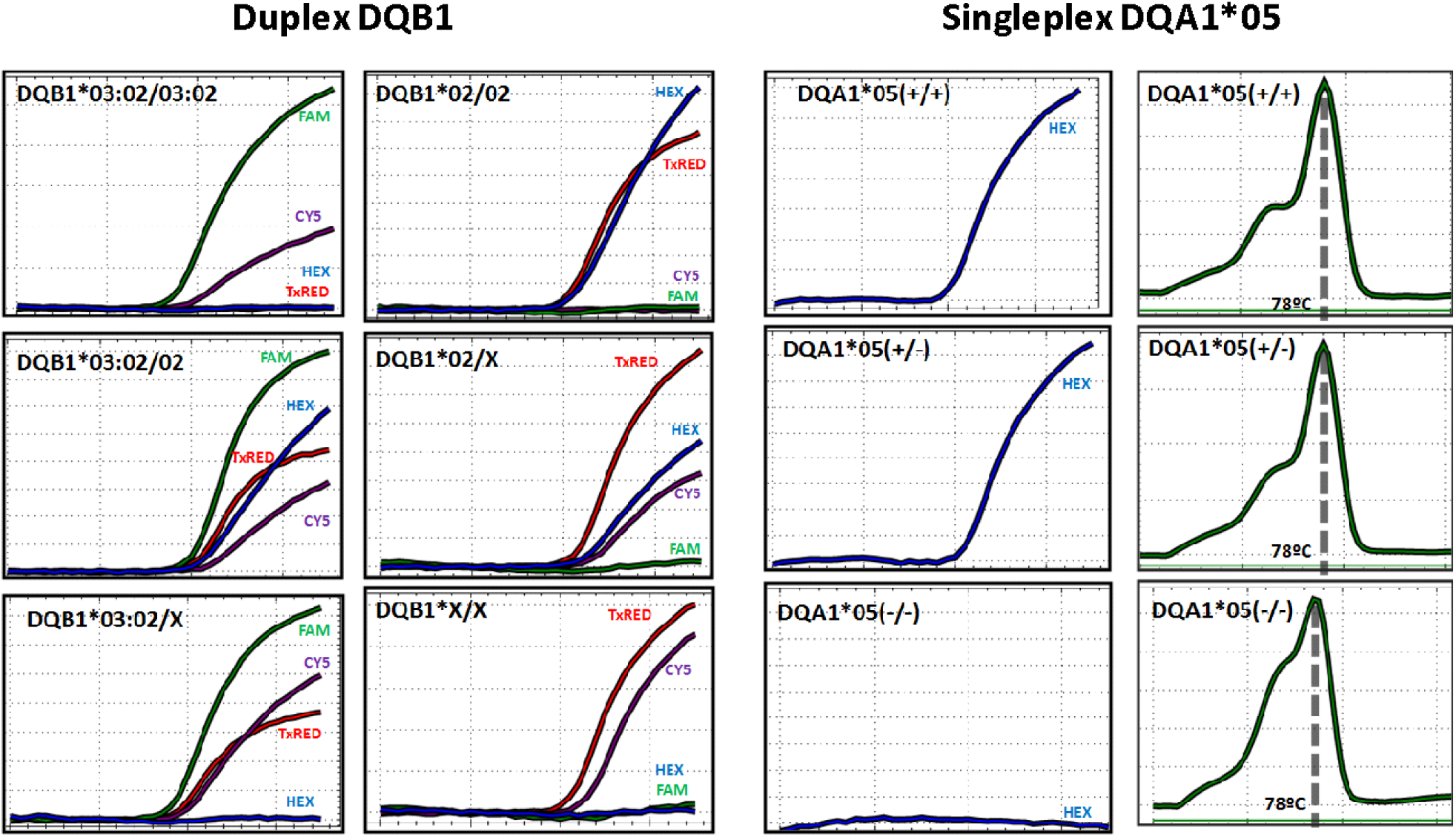
Representative results of direct PCR from saliva. Duplex DQB1 assay. Samples harboring the six possible genotypes derived from three DQB1 alleles (*03:02, *02, and *X) were genotyped using the DQB1 duplex reaction as described. **Singleplex DQA1*05 assay**. Samples carrying DQA1*05, either in homozygosity (+/+) or heterozygosity (+/−), as well as samples lacking the allele (−/−), were genotyped. Amplification curves (acquired at 95ºC, HEX channel) and amplicon melting curves (FAM channel) are shown in the left and right panels, respectively.

